# Utilising Random Effects Models to Analyse Multiple Mini-Interviews for Prospective Medical Students – From Theory to Practice

**DOI:** 10.1101/2025.05.28.25328486

**Authors:** Chezko Malachi Castro, Emanuele Giorgi, Nicola Phillips, Iain Robinson

## Abstract

**Background:** Multiple-mini-interviews (MMIs) are the most commonly used non-academic assessment tool for British medical school admissions processes. Potential inconsistencies can arise from running MMIs, such as differing marking standards among interviewers and stations with varying levels of difficulty.

**Methods:** With the aim of analysing MMI data, the cumulative probit mixed model was deployed, which accounts for variation inherent to MMI scores – both external factors, such as interviewer stringency and station complexity, as well as the factor of interest – candidates’ true performance at interview. With the secondary aim of making this methodology more accessible to non-statistical experts, we developed a user-friendly application using R Shiny. The app was created to standardise MMI scores and generate feedback for interviewers without requiring prior knowledge of programming.

**Results:** Multiple mini-interview data from Lancaster Medical School were analysed for the academic year 2022-2023. Applicant ability (n = 352) contributed to 66.84% of the total variance in MMI scores. Notably, interviewers (n = 83) contributed a considerable proportion of variance (26.50%). Station difficulty (n = 9) had a minor impact on variance (5.81%), with inter-station reliability being acceptable based on the Cronbach’s Alpha value (α = 0. 7072).

**Conclusion:** The current method provides a statistically robust approach towards the analysis of MMI scores. In addition, its companion application can be used by admissions staff to communicate feedback to interviewers on their scoring patterns and identify stations that are better at discriminating applicants’ ability. The illustrated approach can be adapted for use by other medical institutions that use MMI scores to rank candidates during their admissions processes. Future research could investigate how to extend the proposed modelling approach to address applicants’ background factors, such as socio-economic status, for more comprehensive analysis and more equitable offer allocation.

## Introduction

Traditionally, the medical school admissions process has greatly emphasised academic ability, gradually shifting towards a more holistic approach of assessing applicants’ attributes, attitudes, and skills. Within the British context, this has been accompanied in recent years by plans to increase medical school cohort sizes as per the National Health Service’s Long Term Workforce Plan.^1^ To balance the need to meet escalating demand for doctors in an increasingly pressured healthcare system with the justifiable, yet expensive cost of training medical students, it falls to medical schools to develop robust assessment methods by which applicants deemed most “suitable” for medicine can be selected.

Historically, selection for medical school candidates utilised the traditional panel interview format. This has been replaced by a majority of medical schools by the MMI format, or multiple-mini interviews, which alongside markers of academic attainment (e.g. predicted grades) and performance in medical aptitude tests, such as the University Clinical Aptitude Test (UCAT) or the now-defunct BioMedical Admissions Test (BMAT), provide the basis for making offers. First used in medical admissions processes at McMaster University in 2004, MMIs consist of a series of short objective structured clinical examination (OSCE)-style stations that allow assessment of various topics relevant to medicine, such as communication skills and knowledge of healthcare issues.^2^ Indeed, current recommendations from the Medical Schools Council favour the use of MMIs alongside academic attainment and aptitude tests for their selection processes, as stated in the Selecting for Excellence report.^3^

Within the wider literature, there is increasing evidence that MMIs have predictive validity for future academic performance – such as in national licensing examinations and OSCEs.^4-6^ However, a common flaw affecting the reliability of MMIs is a lack of control in the variation of marking behaviours between interviewers (henceforth referred to as inter-rater variability).

Most medical schools conduct MMIs over several days, with simultaneous circuits, and with a wide array of interviewer backgrounds, such as academic staff, clinicians, or medical students, each with their own unconscious scoring behaviours and candidate preferences.^7,8^ As proposed by Myford and Wolfe in a 2004 paper, effects of raters (interviewers) on score can be categorised into five groups: leniency/severity, central tendency, randomness (inconsistency), the halo effect, and differential leniency/severity (i.e. restriction of range).^9^ In addition to these behaviours is the concept of “contrast effects”, where the assessed performance of one candidate is affected by the previous candidates’ performance, which has been observed in similar assessment formats.^10^ Through such mechanisms, interviewers may significantly contribute to the unexplained variance in MMIs and hence decrease the reliability of the admissions process if not properly addressed.

Several papers have utilised the multi-factorial Rasch model (MFRM) or generalisability theory (G theory) in their analysis of MMI to account for variation in MMI scores.^11-13^ MFRMs provide rich information regarding the interactions between interviewer, station, and interviewee through the use of facets in analysing MMI data. Whilst MFRMs may be statistically efficient with increasingly large pools of MMI candidates (it requires a large number of observations to reliably estimate parameters), it can become time-intensive due to the need for repeated iterations in fitting observations to the model. Furthermore, the analysis and interpretation of assessment data using MFRMs require that admissions staff be proficient in the relevant statistical software program.^14^

To resolve the tension between statistical rigour and ease of use, we pursue the two following objectives: 1) To develop a statistically efficient and robust modelling approach to estimate applicants’ abilities while accounting for additional variation in MMI scores introduced by interviewers’ marking severity, station difficulty, and day of interview; and 2) To illustrate a user-friendly interface that allows for ease of use of the above method by practitioners who do not have formal training in statistics.

In meeting our primary objective, we model the MMI scores using cumulative probit mixed models (CPMM)^15(p122-123)^ by utilising random effects to model variation in candidates’ MMI scores, alongside the three sources of residual variation – interviewer, station, and day of interview. We argue that this modelling framework has two main advantages over standard approaches based on MRFMs. The first is that it is statistically more efficient as it exploits the information provided by the ordering of the scores, thus requiring the estimation of a smaller number of model parameters; in addition, the use of random effects also allows us to borrow strength of information across the data which can help reduce uncertainty to estimate each candidate’s performance.

To improve accessibility of this approach, we propose a digital application format as a solution for conducting MMI station scoring analysis. By utilizing a user-friendly interface created with Shiny, an R-based application framework^16^, both standards can be met. Previous studies have highlighted the use of Shiny applications to simplify statistical modelling in educational and psychometric assessments,^17^ but this study is the first to apply a Shiny application within the context of medical education.

## Methods

### Selection of candidates for interview

The selection process for the 2022-2023 admissions cycle (2023 entry) at Lancaster Medical School consisted of the following stages: candidate’s UCAS applications were first screened against the academic entry requirements, then ranked according to their BMAT test results, with the higher-scoring applicants shortlisted for interview. Overseas and Gateway Year applicants were processed separately and were excluded from this analysis.

Interviews were conducted online, over a total of seven days, with duplicate MMI circuits held on each day across two months. Applicants who failed to attend the interview were excluded from the data set, leaving a final sample set of 352 interviewed applicants included in the final analysis.

### Interviewer selection

In total, 83 interviewers were selected from various stakeholder groups: academic staff from Lancaster Medical School, clinicians involved in medical education from primary care and hospital settings, medical students, and members of our patients and public representatives group. Each interviewer was required to complete interviewer training, which comprised information regarding the purpose of MMI, skills and attributes assessed during the stations, instructions on using the rating scales, conducting the mini-interview, and unconscious bias.

To reduce inter-rater variability, interviewer backgrounds were matched with relevant MMI stations. For example, student interviewers were placed exclusively at the student-led station (where students had also developed the task, questions, and marking criteria).

### Format of MMI/Scoring Criteria

The MMI consisted of nine assessed stations, including one standalone 20-minute group discussion station (station C1) and eight 5-minute MMI stations (stations A1-4 and B2, B3, B4, B6). Each station was assessed by one interviewer. In addition to the assessed stations, the MMI circuit also contained two 5 –minute preparation stations, where the applicant is given a task to complete prior to the assessed station, two break stations and an administration station. This time was chosen to minimise the overall assessment period while maintaining the ability to differentiate between applicants and test reliability.

In each MMI station, interviewers were asked to score candidates in three domains, using a five-point scale, creating a maximum scoring range from 3-15. These three domains mainly consisted of two skill domains and a global domain indicating the interviewer’s overall impression of a candidate’s suitability to study medicine given their performance in that station. Within the scale, the marking criteria associated with the mid-point score (3) represented the standard expected of a medical student at the point of entry. At this score, a student would be likely able to cope with the demands of a medical degree. Likewise, the high-point score (5) corresponded to an outstanding applicant whose performance was above what was expected at the point of entry, and the low-point score (1) demonstrated that an applicant’s performance was below expectations and may struggle with the demands of the course.

All stations were trialled before implementation by first-year medical students and senior interviewers to ensure the clarity of instructions and appropriateness of marking criteria. Topics assessed at different stations were taken from various content domains, testing generic attributes desirable for medical school, such as communication skills and knowledge of medical ethics principles. Scores were recorded for analysis in an Excel file, and identifiable candidate information was anonymised by use of candidate number and UCAS ID.

### Statistical analysis and application development

For the analysis of MMI scores, we developed a cumulative probit mixed model (CPMM)^15(p122-123)^ to describe the relationship between observed scores, candidates, interviewers, day of the interview, and station difficulty.

Let *Y*_*ijkt*_ represent the observed score of a particular candidate (*i*) from a particular interviewer (*j*) from a particular station (*k*) on a particular day (*t*). We assume *Y*_*ijkt*_ represent the discretised version of a continuous latent variable *Y\**_*ijkt*_, which is derived from candidate ability (*S*_*i*_), interviewer severity (*T*_*j*_), station difficulty (*U*_*k*_), and effects based on the day of the interview (*V*_*t*_). Additionally, we assume that *Y\**_*ijkt*_, conditionally on + *S*_*i*_, *T*_*j*_, *U*_*k*_, and *V*_*t*_ is a Gaussian distribution, with a mean of µ + *S*_*i*_ + *T*_*j*_ +*U*_*k*_ + *V*_*t*_, where µ = overall average student performance.

Therefore, we can write that:

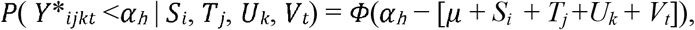

Where *α*_*h*_ are the thresholds such that an observed MMI score *Y*_*ijkt*_ satisfies

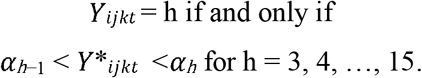

We also assume that *S*_*i*_, *T*_*j*_, *U*_*k*_, and *V*_*t*_ are each independent, zero-mean Gaussian distributions, with respective variances of 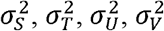.

The objective in utilising this model is to make inferences based on S_i_, which represents the random effect of candidate ability (how much of an impact a candidate makes on the overall interview scores independent of interviewers, station, or days). Under ideal circumstances, the respective contributions of these external factors should be T_j_=U_k_=V_t_=0 for all *j, k*, and *t*. This situation is unlikely to be the case in real-world settings, and so these variables must also be estimated via maximum-likelihood estimation to understand their relative contribution to total variation in MMI scores. Said variables are also scaled according to the variation within the uploaded dataset, such that the effect of an individual candidate’s ability on their MMI scores is generalisable to all theoretical cohorts using the same stations.

The analysis was undertaken by developing a Shiny R application^16^ within the R program (4.3.0),^18^ using the package “ordinal” to fit the CPMM to the MMI data.^19^ The Shiny package was used in order to create a digital interface by which admissions staff could upload a spreadsheet of candidate MMI scores and receive a ranked list of candidates based on the S_i_. Additional functions were also added to the app for ease of analysis, displaying T_j_ (interviewer severity/leniency) and U_k_ (station difficulty/easiness), as well as simple graphical feedback for interviewers.

## Results

### Analysis of MMI scores

A total of 352 candidates’ MMI scores were analysed through the CPMM method, utilising their individual random effects value to create a ranked list of candidates for offers.

Candidates who performed above average at interview (independent of interviewer or station-induced effects) would have positive random effects values, and vice versa, with the magnitude of their (*S*_*i*_) value representing to what extent a candidate’s ability at interview impacted their MMI scores in standard deviations from the mean. Based on the histograms of estimated random effects values of candidates and interviewers (Figures 1 and 2), candidate ability and interviewer severity appear to be normally distributed, confirming our prior assumptions required for the CPMM to hold.

**Figures 1 and 2:**
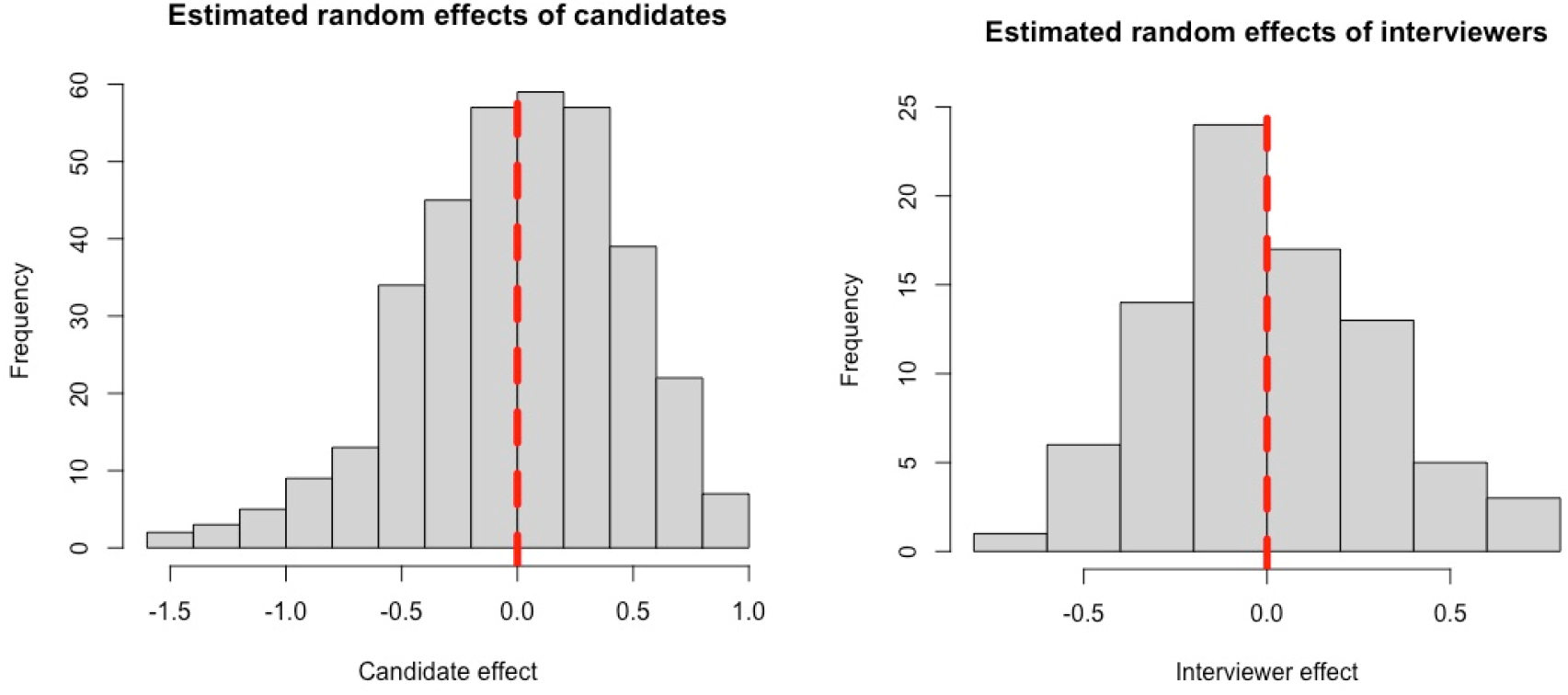

Cronbach’s alpha was calculated to be acceptable (α= 0.7072). In line with expectations, candidate ability contributed the most to variation (66.8%); notably, interviewers marking behaviours explained a significant proportion of variance in scores (26.5%). Differences due to station difficulty (5.81%) or day (0.85 %) had little effect on MMI scores (Table 1).

**Table 1:** Estimated variances of random effects and contribution to total variance within MMI scores.

| Variables | Standard deviation of random effects | Variance of random effects | Total contribution to variance (total variance = 0.4390) |
| --- | --- | --- | --- |
| Candidate ability<br>(n = 352) | 0.5417 | 0.2934 | 66.84% |
| Interviewer marking behaviours<br>(n = 83) | 0.3411 | 0.1164 | 26.50% |
| Station difficulty<br>(n = 9) | 0.1597 | 0.0255 | 5.81% |
| Day-induced effect<br>(n = 7) | 0.0610 | 0.0037 | 0.85% |

Particular stations were identified on analysis as having random effects greater than one, or less than minus one – for example, station B6 and station B7, with random effect values of 1.23 and -1.10 respectively (Table 2 and Figure 3). These stations were kept in the analysis due to the necessity of the attributes tested at each station. The high positive effect of B6 suggested that candidates likely found the content of the station easier, which thus had a positive impact on candidate scores overall, whilst station B7 was particularly difficult and negatively impacted candidate scores.

**Table 2:** MMI stations and the corresponding random effects values. Different station IDs are presented, each assessing different topics.

| Station ID | Station Random Effect ($U_k$) |
| --- | --- |
| A1 | 1.0454 |
| A2 | -0.6506 |
| A3 | 0.0618 |
| A4 | -1.0573 |
| B2 | -0.1522 |
| B3 | -0.02 |
| B6 | 1.2071 |
| B7 | -1.0749 |
| C1 | 0.6336 |

**Figure 3:**
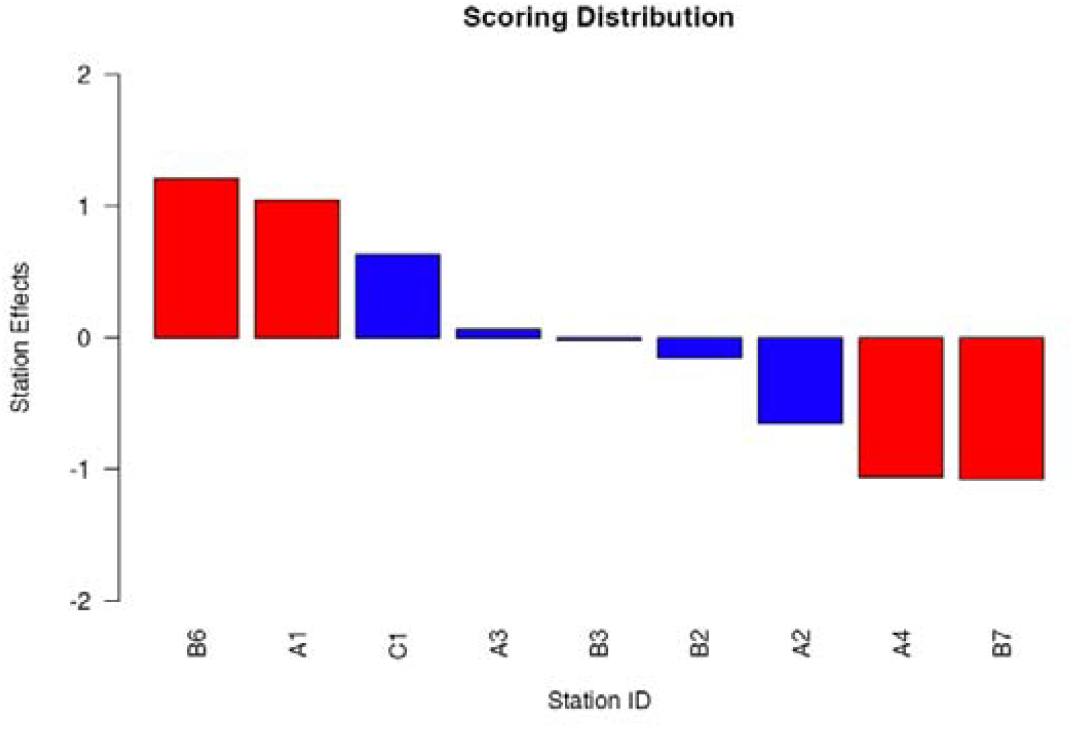
Bar chart showing the random effects of MMI stations. Note the stations A1, A4, B6, and B7, coloured red, indicating values one standard deviation above and below a random effect of zero.

### Application use

The Shiny application was successfully trialled and required little computing time to fit the cumulative probit mixed model to the data. Upon completion, it was able to generate a ranked list of interview candidates by performance, as well as station and interviewer feedback.

Simple descriptive statistics were given to interviewers based on their impact on MMI scores. The operational manual will be attached to this paper as supplementary material.

Images 1 and 2 display screenshots of the Shiny application used for the MMI score analysis. Image 1 depicts the candidate ranking table generated by the application after the relevant file was uploaded and analysed. As stated previously, candidates with high random effects values were determined to have high performance at interview. Image 2 is a screenshot from the same application, showing feedback given to an individual interviewer.

**Images 1 and 2:**
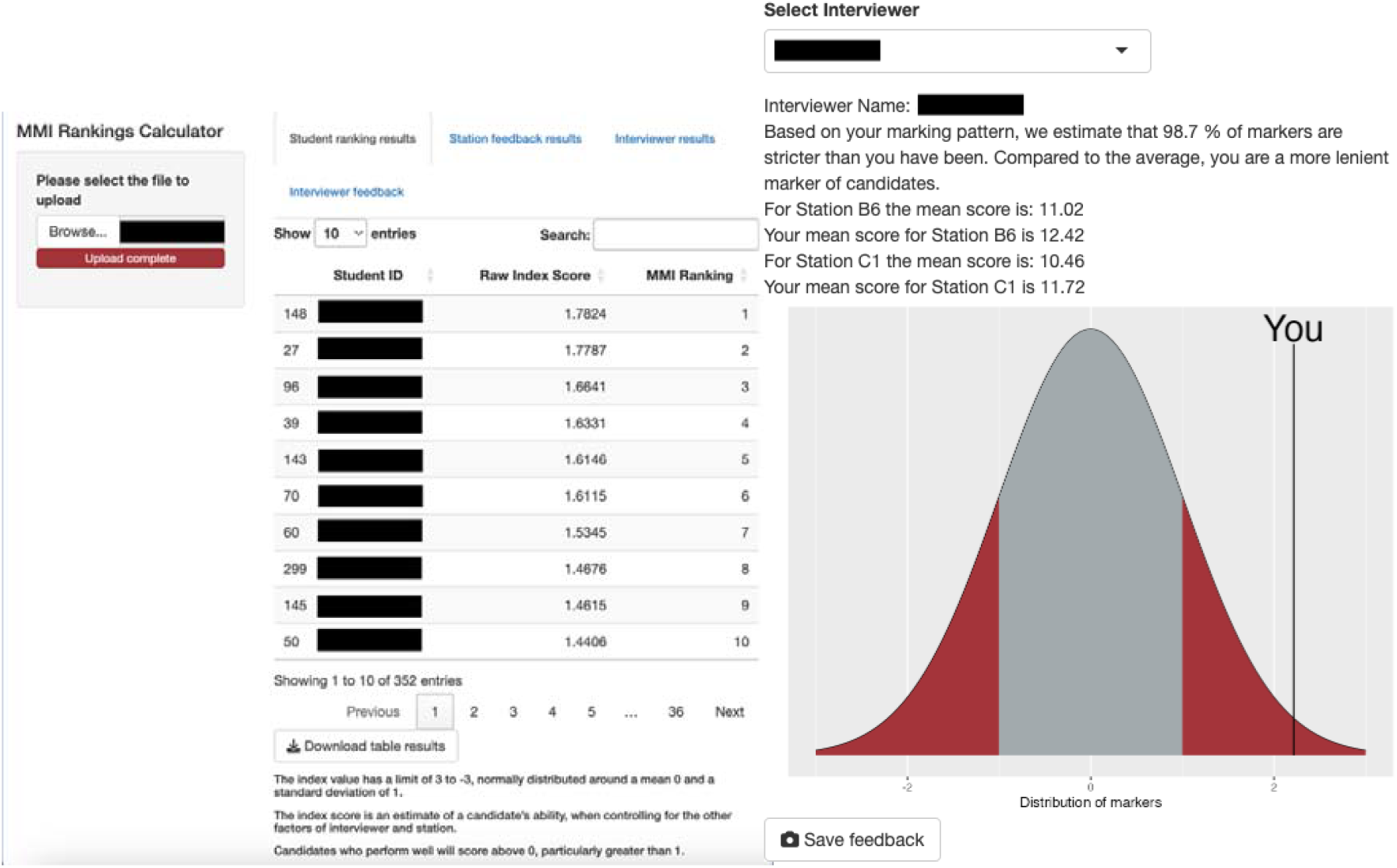

## Discussion

The CPMM approach is successful at creating a sound basis for an offer allocation system, as it partitions effects attributable to specific variables, including external variables that can increase variation in candidates’ performance at interview. We found that candidates’ ability at interview explained the large majority of variance in MMI scores. However, this is out of keeping from the wider body of literature, which utilises MFRMs or G theory in their analyses. Such studies show more modest attributable variance – Roberts et al.^20^ and Till et al.^21^ report an attributable percentage to student ability of 19.1% and 16.01%, respectively. For both of these papers, the total attributable proportion of variance (e.g. to candidates or interviewers) by the Rasch model is less than the proportion of unexplained variance. This discrepancy in results between MFRMs and CPMMs is purely methodological due to differences in calculating attributable variance for each component of the MMI.

The CPMM approach also allows for the scrutiny of MMI station design. Whilst this variable did not take a large proportion of variance, the probit model still allowed for the identification of stations that tended towards the extremes of difficulty for the average medical applicant cohort. To improve future analyses, marks per station could be subdivided into the assessed domains, such that rather than stations, “topics assessed” would be the random effects variable. Additionally, this would help to unmask measurement error (for example, if the global performance mark produced undue weighting on candidates’ MMI scores).

In quantifying the external random effects variables, this method can be simultaneously deployed to identify problematic interviewer patterns – in particular, leniency/severity. Within the 2022 cohort, over a quarter of explainable variance was due to marking behaviour – in keeping with the extant literature, as this variable was found to be the second-largest source of explainable variance in MMI scores in papers using the MFRM method.^20,21^ Similarly, the inter-station variability (Cronbach’s alpha) is also in keeping with other studies utilising MFRM.^12^ Whilst a level of 0.7+ is regarded as acceptable, it must be noted that higher values of Cronbach’s alpha are unlikely to be expected due to the range of topics assessed within MMI stations at Lancaster.

In contrast to MFRMs, the interpretation of CPMM requires less statistical knowledge, making it feasible for use in admissions processes. Furthermore, it is more statistically efficient, as there are fewer parameters to estimate. However, further improvements must still be made to improve its capabilities in capturing rater effects. From the aforementioned rater effects^9^, the current specification of the cumulative probit mixed model explicitly accounts for leniency/severity. It could be further improved to address differential leniency/severity through calculating rater-candidate interactions. It would also be worthwhile to account the effects that various facets of a candidate’s background, such as socioeconomic status, might have on their MMI scores. Although the literature produces mixed results as to whether such an effect exists,^22^ the CPMM is flexible in terms of what additional sources of variation it could account for, potentially allowing for fairer offer allocation.

Based on our institution’s experience of MMI analysis using cumulative probit mixed models over the last few years, we have found this approach to be a statistically robust and accessible tool in our medical admissions process. Areas for further research include further use of the model in scrutinising the effects of applicant demographics and aberrant marking behaviour detection on MMI scores, as well as application of the model within other institutions.

## Conclusion

By utilising a CPMM approach in the analysis of MMI scores, we present a statistically robust method of selectively capturing candidates’ ability to perform at interview, that, through a digital application, allows for ease of analysis within admissions processes by creating a basis for a ranked offer system. This modelling method allows for the detection of interviewer severity/leniency and station easiness/difficulty through its estimation of random effects variables which would otherwise impact MMI scores. The majority of variance in MMI scores was primarily due to variation in candidate ability, with a smaller proportion attributable to interviewer marking behaviour. Further research should focus on extending the CPMM models by incorporating interviewer-specific information, such as their level of training and historical marking behaviour, to enhance the robustness of standardization. By refining the model to account for these factors, the approach can better mitigate variations in interviewer severity or leniency, further strengthening the reliability of MMI scoring. The proposed analytical approach to MMIs has enabled admissions staff to provide informed choices regarding offers by limiting the amount of undue influence external variables have on a candidate’s MMI score.

## Supporting information

code for the application

user manual

## Statements and Declarations

### Ethical considerations

Ethical approval has been granted by the Faculty of Health & Medicine Research Ethics Committee at Lancaster University.

### Consent to participate

Not applicable.

### Consent for publication

Not applicable.

### Declaration of conflicting interest

The authors declared no potential conflicts of interest with respect to the research, authorship, and/or publication of this article.

### Funding statement

The author(s) received no financial support for the research, authorship, and/or publication of this article.

### Data availability

The data used in the analysis will not be made available to the public due to its confidential nature and is protected under copyright. The code for the application will be publicly available and can be accessed digitally in the supplementary material alongside the user manual.

## Notes

### Competing Interest Statement

The authors have declared no competing interest.

### Funding Statement

This study did not receive any funding.

### Author Declarations

The Faculty of Health and Medicine Research Ethics Committee of Lancaster University gave ethical approval.

