## Supplementary material for "Utilising Random Effects Models to Analyse Multiple Mini-Interviews for Prospective Medical Students – From Theory to Practice": code for the application

```
# Loading necessary libraries
library(shiny)
library(ordinal)
library(DT)
library(readxl)
library(dplyr)
library(shinyscreenshot)
library(ggplot2)
library(writexl)




# Code for UI

ui <- fluidPage(
  tags$head(tags$style(".progress-bar{background-color:#A4343A;}")),
  fluidRow(
    column(4,h4("MMI Rankings Calculator"),
           wellPanel(
             fileInput('mmi_file', 'Please select the file to upload',
                       accept = c(
                         'text/csv',
                         'text/comma-separated-values',
                         'text/tab-separated-values',
                         'text/plain',
                         '.csv',
                         '.tsv',
                         '.xlsx'
                       )
             )
           )

    ),
    mainPanel(
      tabsetPanel(type = "tabs",
                  tabPanel(h6("Student ranking results"),
                           dataTableOutput("ranking_students"),
                           downloadButton("download", "Download table results"),
                           h6("The index value has a limit of 3 to -3, normally
distributed around a mean 0 and a standard deviation of 1."),
                           h6("The index score is an estimate of a candidate's ability,
when controlling for the other factors of interviewer and station."),
                           h6("Candidates who perform well will score above 0,
particularly greater than 1."),
                           plotOutput("studentgraph")),
                  tabPanel(h6("Station feedback results"),
                           dataTableOutput("ranking_stations"),
                           h6("The index value has a limit of 3 to -3, normally
distributed around a mean 0 and a standard deviation of 1."),
                           h6("The index measures the impact a station has on
discriminating candidates who do poorly versus those who do well."),
                           h6("If a station has a score above 1, it is likely that the
station is particularly easy (that candidates tend to score well overall); a score below
-1 also suggests poor discrimination of candidate performance (that most students tend to
score poorly)."),
                           h6("Stations at the extremes (>1 and <-1) should be assessed
for difficulty or necessity."),
                           plotOutput("station_plot")),
                  tabPanel(h6("Interviewer results"),
                           dataTableOutput("ranking_interviewers"),
                           h6("The index value has a limit of 3 to -3, normally
distributed around a mean 0 and a standard deviation of 1."),
                           h6("The index measures the impact an interviewer has on
discriminating candidates who do poorly versus those who do well."),
                           h6("The ideal range for an interviewer's impact is 1 < x < -1.
"),
                           h6("If an interviewer's index value is less than -1, it
```

```
suggests that an interviewer has an over-excessive discrimination between students - that
candidates with higher ability are scored higher and those with lower ability are scored
lower. A score greater than 1 suggests that an interviewer may be relatively lenient with
their marking criteria, giving most people a similar station score regardless of the
student ability.")),
                  tabPanel(h6("Interviewer feedback"),
                           uiOutput("interviewer_names"),
                           uiOutput("interviewer_feedback"),
                           plotOutput("interviewer_feedback_graph"),
                           screenshotButton(label = "Save feedback", selector = ".tab-
content")))
    )



  ))




# The shiny app server command
server <- function(input, output, session) {

  df = reactive({
    req(input$mmi_file)
    uploaded_data <- read_xlsx(input$mmi_file$datapath)

    colnames(uploaded_data) <- c("day", "number", "station", "interviewer", "score",
"student")
    uploaded_data$score <- factor(uploaded_data$score)

    return(uploaded_data)
  })

  fittedModel <- reactive({
    req(df())
    clmm(score ~ 1 + (1 | station) + (1 | interviewer) + (1 | student) + (1 | day),
         data = df(), link = "probit", threshold = "flexible")
  })

  ranking_df <- reactiveVal(NULL)
  observe({
    req(fittedModel())

    re.hat <- ranef(fittedModel())
    std.hat <- fittedModel()$ST

    latent.score <- as.numeric(re.hat$student[,1]) / as.numeric(std.hat$student)
    ranking.score <- sort.list(latent.score, decreasing = TRUE)
    student.effect <- data.frame('Student ID' = rownames(re.hat$student),
                                 'Student ranking index ' = round(latent.score, 4))

    ranked_df <- student.effect[ranking.score, ]
    ranked_df$'Student Ranking' <- sort(ranking.score)
    colnames(ranked_df) <- c("Student ID", "Raw Index Score", "MMI Ranking")

    ranking_df(ranked_df)
  })

  output$ranking_students <- renderDataTable({
    req(ranking_df())
    ranking_df()
  })

  output$download <- downloadHandler(
    filename = function() {
      paste("MMI_Results", Sys.Date(), ".xlsx", sep = "_")
    },
    content = function(file) {
      write_xlsx(ranking_df(), file)
```

```
    }
  )


  output$studentgraph <- renderPlot({
    req(fittedModel())


    p2 <- ggplot(data = data.frame(x = c(-3, 3)), aes(x)) +
      stat_function(fun = dnorm, n = 101, args = list(mean = 0, sd = 1)) + ylab("") +
xlab("Distribution of students") +
      scale_y_continuous(breaks = NULL) +
      stat_function(fun = dnorm,
                    xlim = c(-3,-1),
                    geom = "area", fill = "red") +
      stat_function(fun = dnorm,
                    xlim = c(1,3),
                    geom = "area", fill = "green") +
      stat_function(fun = dnorm,
                    xlim = c(-1,1),
                    geom = "area", fill = "gold")
    p2

  })

  output$ranking_stations <- renderDataTable({
    #ranks the stations
    req(fittedModel())

    re.hat <- ranef(fittedModel())
    std.hat <- fittedModel()$ST

    latent.station.score <- as.numeric(re.hat$station[,1]) / as.numeric(std.hat$station)
    station.ranking.score <- sort.list(latent.station.score, decreasing = TRUE)
    station.effect <- data.frame('Station ID' = rownames(re.hat$station),
                                 'Station  score' = round(latent.station.score, 4))

    station.ranking_df <- station.effect[station.ranking.score, ]
    station.ranking_df$'Ranked Station Index Score' <- sort(station.ranking.score)

    colnames(station.ranking_df) <- c("Station ID", "Raw Station Score", "Station
Ranking")

    return(station.ranking_df)

  })

  output$station_plot <- renderPlot({
    #makes a plot of the station rankings
    req(fittedModel())

    re.hat <- ranef(fittedModel())
    std.hat <- fittedModel()$ST

    latent.station.score <- as.numeric(re.hat$station[,1]) / as.numeric(std.hat$station)
    station.ranking.score <- sort.list(latent.station.score, decreasing = TRUE)
    station.effect <- data.frame('Station ID' = rownames(re.hat$station),
                                 'Station score' = round(latent.station.score, 4))

    station.ranking_df <- station.effect[station.ranking.score, ]
    station.ranking_df$'Ranked Station Index Score' <- sort(station.ranking.score)

    colnames(station.ranking_df) <- c("Station ID", "Station Effect", "Station Ranking")


    barplot(height = station.ranking_df$"Station Effect",
            main = 'Scoring Distribution',
            xlab = 'Station ID', ylab = "Station Effects", ylim = c(-2,2),
            names.arg = station.ranking_df$"Station ID", las = 2,
            col = case_when(
```

```
              station.ranking_df$"Station Effect" > 1 ~ "red",
              station.ranking_df$"Station Effect" < -1 ~ "red",
              .default = "blue")


    )
    #    p3 <- ggplot(data = station.ranking_df, aes(x = 
reorder(station.ranking_df$"Station ID",-station.ranking_df$'Raw Station Score'),
    #                                             y = station.ranking_df$'Raw Station 
Score',
    #                                              fill = station.ranking_df$'Raw Station 
Score' > 1 || station.ranking_df$'Raw Station Score' < -1)) + geom_bar(stat = "identity") 
+
    #   scale_fill_manual(values = c("red","green"), labels=c('TRUE'='Out of 
range','FALSE'='Within range'))
    #   p3
  })

  output$ranking_interviewers <- renderDataTable({
    #makes a ranking for the interviewers
    req(fittedModel())

    re.hat <- ranef(fittedModel())
    std.hat <- fittedModel()$ST

    interviewer.latent.score <- as.numeric(re.hat$interviewer[,1]) / 
as.numeric(std.hat$interviewer)
    interviewer.ranking.score <- sort.list(interviewer.latent.score, decreasing = TRUE)
    interviewer.effect <- data.frame('Interviewer Name' = rownames(re.hat$interviewer), 
                                     'Interviewer latent score' = 
round(interviewer.latent.score, 4))

    interviewer.ranking_df <- interviewer.effect[interviewer.ranking.score, ]
    interviewer.ranking_df$'Interviewer Ranking Index Score' <- 
sort(interviewer.ranking.score)

    colnames(interviewer.ranking_df) <- c("Interviewer Name", "Raw Index Score", 
"Interviewer Ranking")

    return(interviewer.ranking_df)
  })

  output$interviewer_names <- renderUI({
    #for choosing the interviewer name!
    req(fittedModel())
    selectInput('cols', 'Select Interviewer', choices = df()$interviewer, selected = NULL
)
  })

  output$interviewer_feedback <- renderUI({
    #making the text to be given out as feedback.
    req(fittedModel())
    req(input$cols)
    selected_interviewer <- input$cols

    feedback_text <- paste("Interviewer Name: ", selected_interviewer, HTML("<br>"))

    re.hat <- ranef(fittedModel())
    std.hat <- fittedModel()$ST

    interviewer.latent.score <- as.numeric(re.hat$interviewer[,1]) / 
as.numeric(std.hat$interviewer)
    interviewer.ranking.score <- sort.list(interviewer.latent.score, decreasing = TRUE)
    interviewer.effect <- data.frame('Interviewer Name' = rownames(re.hat$interviewer), 
                                     'Interviewer latent score' = 
round(interviewer.latent.score, 4))
    interviewer.ranking_df <- interviewer.effect[interviewer.ranking.score, ]
    interviewer.ranking_df$'Interviewer Ranking Index Score' <- 
sort(interviewer.ranking.score)
```

```
    feedback_text <- paste("Interviewer Name: ", selected_interviewer, HTML("<br>"))
    interviewer_ranking_score <- interviewer.ranking_df$`Interviewer Ranking Index
Score`[interviewer.ranking_df$`Interviewer.Name` == selected_interviewer]
    interviewer_index <-
interviewer.ranking_df$`Interviewer.latent.score`[interviewer.ranking_df$`Interviewer.Name`
== selected_interviewer]


    interviewernorm <- round((pnorm(interviewer_index, mean = 0, sd = 1) * 100), 1) #sd is
already accounted because it's an index - already standardised

    if (interviewernorm > 84.1) {
      feedback_text <- paste(feedback_text, "Based on your marking pattern, we estimate
that ", interviewernorm, "% of markers are stricter than you have been. Compared to the
average, you are a more lenient marker of candidates.", "<br>")
    } else if (interviewernorm < 15.9) {
      feedback_text <- paste(feedback_text, "Based on your marking pattern, we estimate
that ", interviewernorm, "% of markers are stricter than you have been. Compared to the
average, you are a stricter marker of candidates.", "<br>")
    } else {
      feedback_text <- paste(feedback_text, "Based on your marking pattern, we estimate
that", interviewernorm, "% of markers are stricter than you have been. Your marking
behavior is approximately in line with an average marker.", "<br>")
    }
    # Filtering the Station + Interviewer-Specific Feedback
    data_filtered <- df()[df()$interviewer == selected_interviewer,]
    selected_station <- unique(data_filtered$station)



    for (station in selected_station) {
      data_for_station <- df()[df()$station == station, ]
      data_for_station$score <- as.numeric(as.character(data_for_station$score))


      if (any(is.na(data_for_station$score) | is.nan(data_for_station$score))) {
        feedback_text <- paste(feedback_text, "For Station", station, ", the 'score' value
is missing.", "<br>")
      } else {
        mean_station_score <- mean(data_for_station$score, na.rm = TRUE)
        feedback_text <- paste(feedback_text, "For Station", station, "the mean score is:
", round(mean_station_score, 2), "<br>")

        # Filter data for the specific interviewer at the station
        interviewer_data_for_station <- data_for_station[data_for_station$interviewer ==
selected_interviewer, ]

        if (nrow(interviewer_data_for_station) == 0) {
          feedback_text <- paste(feedback_text, "Error - no score for this interviewer at
Station ", station, "<br>")
        } else {
          interviewer_scores_for_station <- as.numeric(interviewer_data_for_station$score)
          # previously did as.character there
          if (any(is.na(interviewer_scores_for_station) |
is.nan(interviewer_scores_for_station))) {
            feedback_text <- paste(feedback_text, "Missing scores for interviewer at
Station ", station, "<br>")
          } else {
            mean_interviewer_score <- mean(interviewer_scores_for_station, na.rm = TRUE)
#Calculating mean interviewer score
            feedback_text <- paste(feedback_text, "Your mean score for Station", station,
"is", round(mean_interviewer_score, 2), "<br>")

          }
        }
      }
    }

    HTML(feedback_text)
```

```
  })

  output$interviewer_feedback_graph <- renderPlot({
    req(fittedModel())
    req(input$cols)

    selected_interviewer <- input$cols


    re.hat <- ranef(fittedModel())
    std.hat <- fittedModel()$ST

    interviewer.latent.score <- as.numeric(re.hat$interviewer[,1]) /
as.numeric(std.hat$interviewer)
    interviewer.ranking.score <- sort.list(interviewer.latent.score, decreasing = TRUE)
    interviewer.effect <- data.frame('Interviewer Name' = rownames(re.hat$interviewer),
                                     'Interviewer latent score' =
round(interviewer.latent.score, 4))
    interviewer.ranking_df <- interviewer.effect[interviewer.ranking.score, ]
    interviewer.ranking_df$'Interviewer Ranking Index Score' <-
sort(interviewer.ranking.score)

    interviewer_ranking_score <- interviewer.ranking_df$`Interviewer Ranking Index
Score`[interviewer.ranking_df$`Interviewer.Name` == selected_interviewer]
    interviewer_index <-
interviewer.ranking_df$`Interviewer.latent.score`[interviewer.ranking_df$`Interviewer.Name`
== selected_interviewer]



    p1 <- ggplot(data = data.frame(x = c(-3, 3)), aes(x)) +
      stat_function(fun = dnorm, n = 101, args = list(mean = 0, sd = 1)) + ylab("") +
xlab("Distribution of markers") +
      scale_y_continuous(breaks = NULL) +
      stat_function(fun = dnorm,
                    xlim = c(-3,-1),
                    geom = "area", fill = "#A4343A") +
      stat_function(fun = dnorm,
                    xlim = c(1,3),
                    geom = "area", fill = "#A4343A") +
      stat_function(fun = dnorm,
                    xlim = c(-1,1),
                    geom = "area", fill = "#A2AAAD")
    p1 + geom_segment(aes(x = interviewer_index, y = 0, xend = interviewer_index, yend =
0.385)) +
      annotate("text", x=interviewer_index, y = 0.4, label="You", angle=0, size=10,
color="black")




  })




}
# Running it -
shinyApp(ui = ui, server = server)
```
