## Supplementary material for "Utilising Random Effects Models to Analyse Multiple Mini-Interviews for Prospective Medical Students – From Theory to Practice": user manual

**MMI Shiny App User Manual**

Requirements for the App:

1. Open the R file containing the Shiny R app and download the required packages when prompted. Press Run App.
2. If you have successfully opened the app, it should look like the image below.
   To upload the data, click the Browse button (highlighted below) in the top left corner and select the relevant file from your system storage, which should be in .xlsx format.

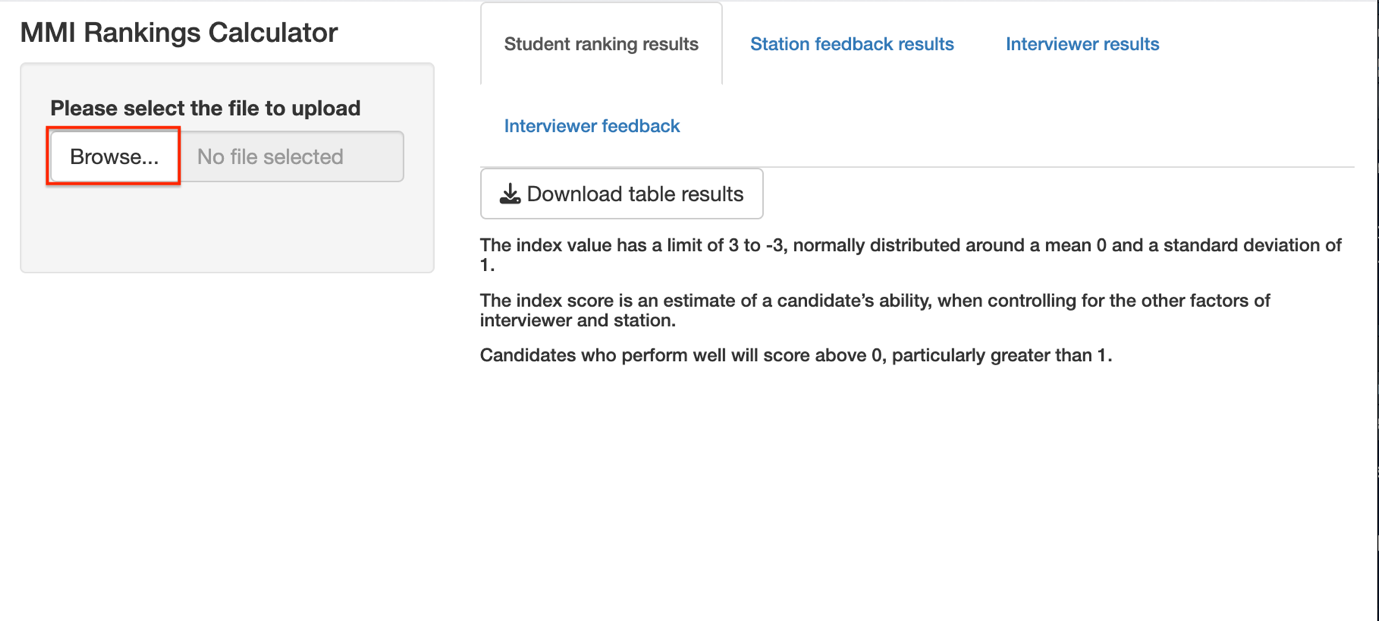

3. Please allow approximately 2-3 minutes’ waiting time for the data to be analysed.
4. From the selected tab “Student ranking results”, a ranking of the students will be displayed once the data has been analysed. You can download the table in .xlsx format by pressing the highlighted button (as seen in the image below). Below the table is a graphical explanation of how to interpret the ranking.
5.
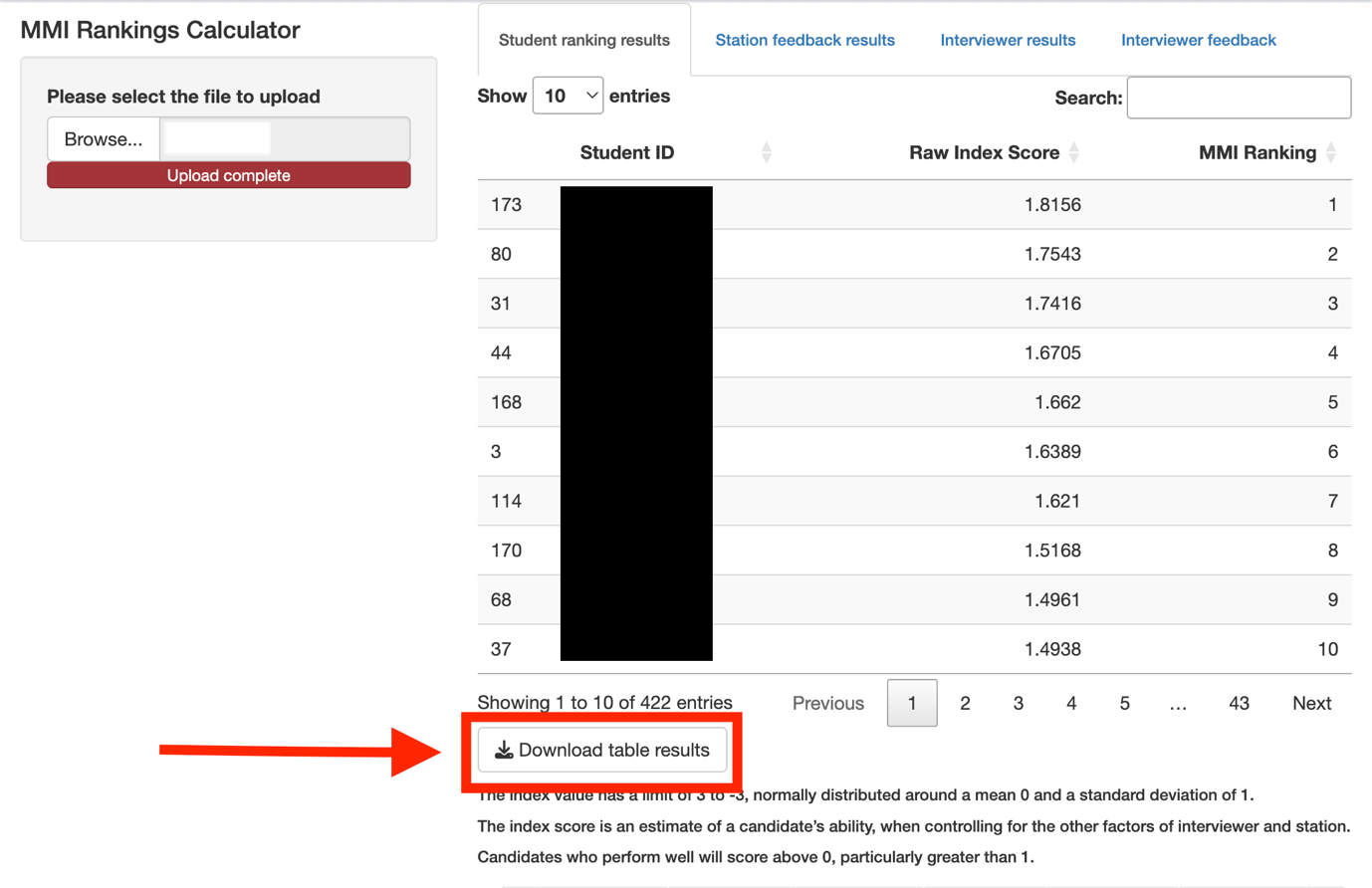
Select the tab named “Station feedback results”. A ranking of the stations will be displayed once the data has been analysed, as shown in the image below.

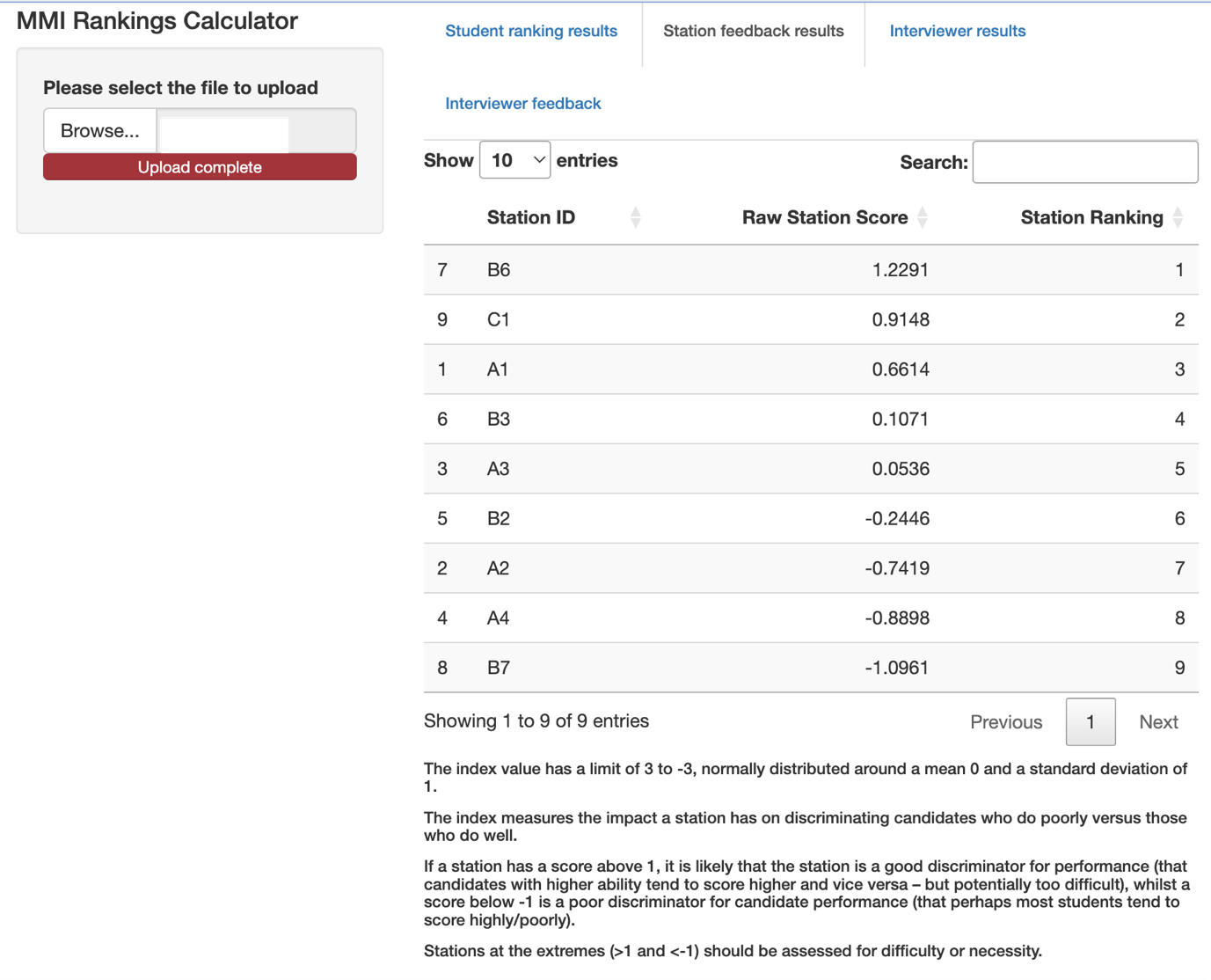

6. Select the tab named “Interviewer results”. A ranking of the interviewers will be displayed once the data has been analysed, with an explanation of the interpretation below it.

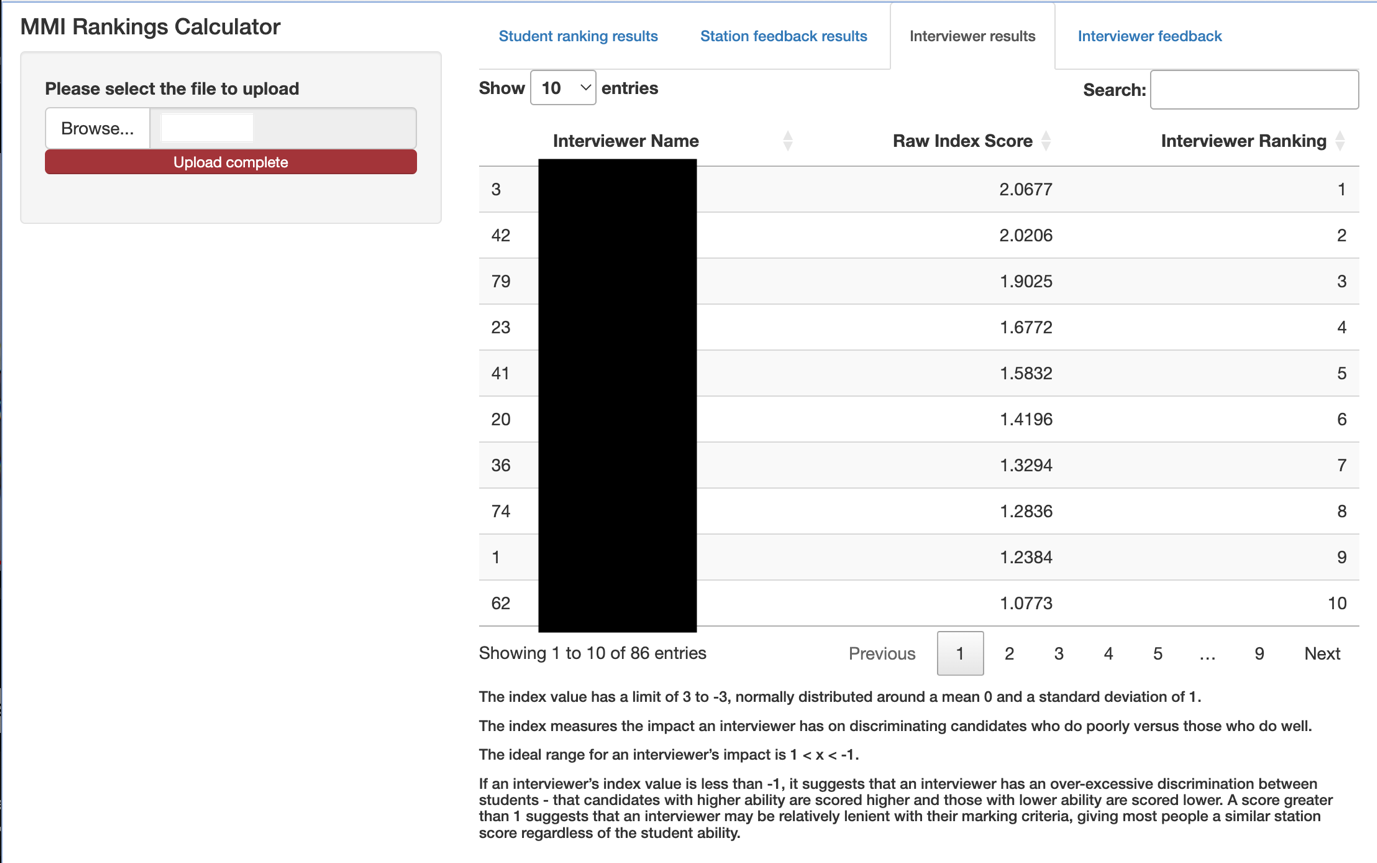

7. Select the tab named “Interviewer feedback.” There will be a multiple-selection box displaying a list of the MMI interviewers. Select the relevant interviewer’s name to display their feedback. There is the option to download the feedback by pressing the Save Feedback button, as shown in the image below.

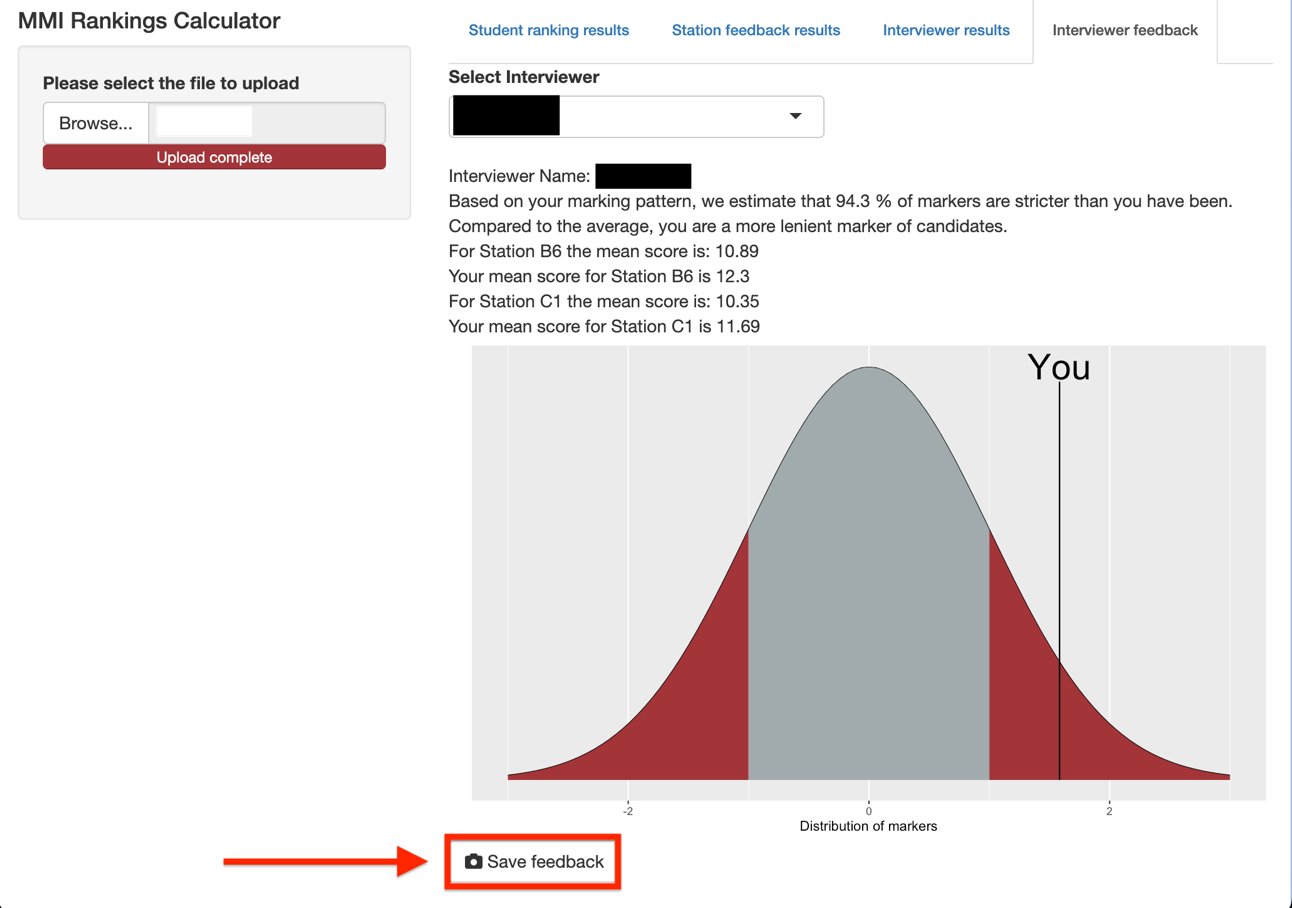

8. When finished, the downloaded feedback file will look like the image below, and can be emailed to the interviewer.

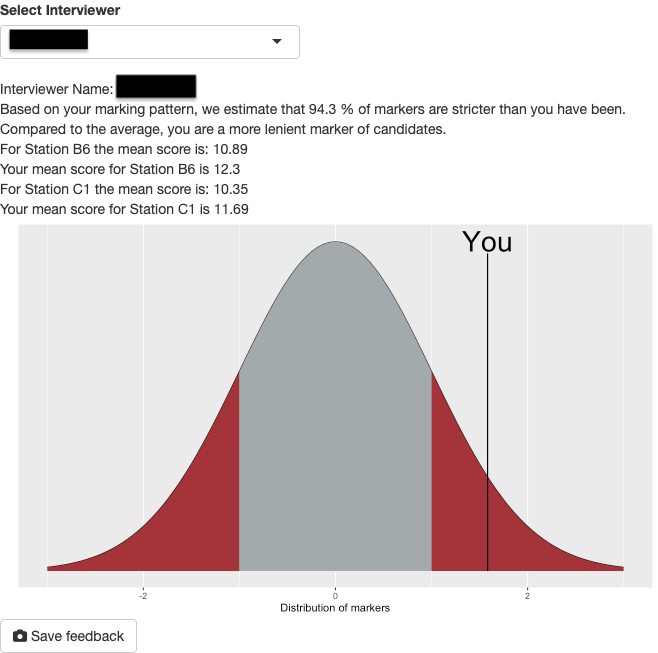
